# Exploring Ophthalmology Content on TikTok

**DOI:** 10.1101/2022.07.07.22277339

**Authors:** Austin Huang, Dagny Zhu

**Affiliations:** Department of Ophthalmology, Baylor College of Medicine, Houston, TX; Hyperspeed LASIK/NVISION Eye Centers, Rowland Heights, CA

**Author notes:** Corresponding Author: Austin Huang, Department of Ophthalmology, Baylor College of Medicine, 1 Baylor Plaza, Houston, Texas 77030.

**Keywords:** TikTok, ophthalmology, social media

## Abstract

TikTok has recently emerged as a rapidly growing global phenomenon that is becoming popular among medical professionals and patients alike. The use of other social media platforms such as Instagram and Twitter by ophthalmologists is well-documented, but the prevalence of ophthalmologists on TikTok is not well understood. We sought to analyze the scope of popular ophthalmology content on TikTok.

## MAIN TEXT

TikTok has recently emerged as a rapidly growing global phenomenon that is becoming popular among medical professionals and patients alike.^1^ The use of other social media platforms such as Instagram and Twitter by ophthalmologists is well-documented,^2,3^ but the prevalence of ophthalmologists on TikTok is not well understood. We sought to analyze the scope of popular ophthalmology content on TikTok.

The TikTok search query was employed to identify the number of views for 37 ophthalmology-related hashtag terms. These terms were utilized in a previous survey of ophthalmology content on Instagram, which combined ophthalmology terms derived from the American Academy of Ophthalmology Intelligent Research in Sight and ophthalmology phrases commonly used by the general public.^3^ The top 10 most-liked videos from each hashtag were then assessed whether they were posted by ophthalmologists or non-ophthalmologists in accordance with their self-posted profile biography and linked social media websites. Veterinary ophthalmic and non-ophthalmology-related posts were excluded from analysis of most-liked posts.

The number of views for all videos associated with each of these 37 hashtags is displayed in Table 1. These videos garnered over 1.452 billion views on TikTok. The most popular hashtags were “LASIK” (234.6 million views), “Blepharoplasty” (183.4 million), “Ophthalmology” (128 million), “EyeSurgery” (119.5 million), “Myopia” (111.7 million), and “DryEye” (105 million). Of the top 10 most-liked videos from each hashtag, 67 out of a total of 370 videos (18.1%) were posted by ophthalmologists.

**Table 1.**
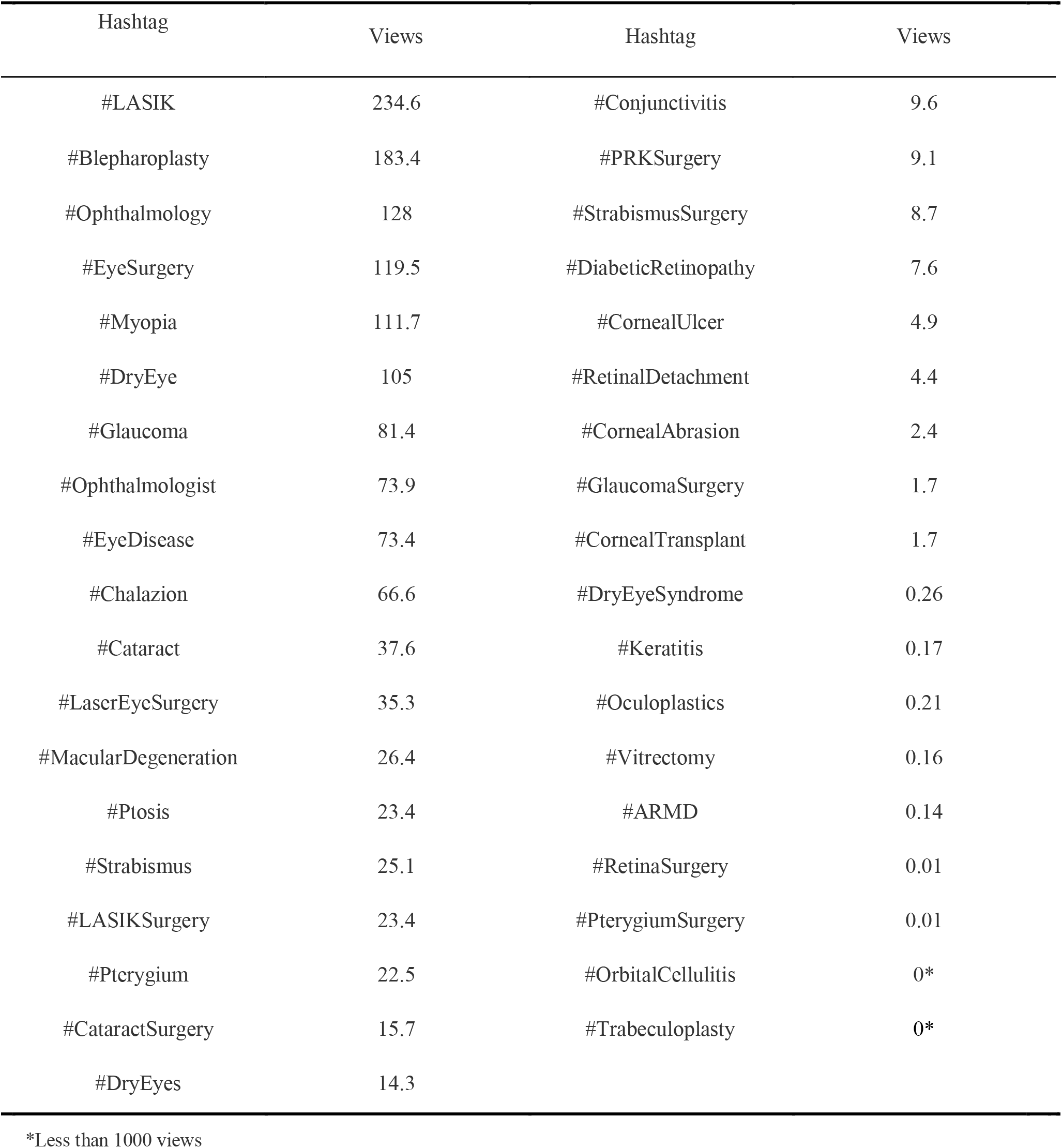
Number of views (in millions) for all ophthalmology-related hashtags searched on TikTok

We determined that only a small minority (18.1%) of the most popular ophthalmology-related videos were created by ophthalmologists. This finding is consistent with TikTok analyses in other fields such as dermatology (15.1%),^1^ but it is a much smaller proportion than ophthalmologist posts on Instagram (35.8%).^3^ This may indicate an underserved niche for ophthalmologists on TikTok, especially given the high level of engagement on ophthalmology-related content by users.

The presence of social media in medicine has grown exponentially over the past decades, and the field of ophthalmology is no exception, from clinicians promoting their practice and personal lives on Twitter and Instagram to academic journals sharing the latest innovations and discoveries.^4^ Augmented by the coronavirus-disease-19 (COVID-19) pandemic and the subsequent shift to a virtual—and now hybrid—society, the importance of social media cannot be overstated. However, the rise of social networking is accompanied by the increase of online dissemination of misinformation, as rapid transmission suffers from a scarcity of timely expert scrutiny.^5^ This is yet another reason why it may be beneficial to have the presence of more ophthalmologists on social media. Limitations to this study include determining if video creators are ophthalmologists or non-ophthalmologists via self-reported social media profiles and websites.

Our results suggest that TikTok is an underused platform for ophthalmologists in comparison to other social media networks, despite the immense popularity of the social networking site over the past couple of years.^1^ The 1.452 billion views accumulated by the videos in the curated list of ophthalmology-related hashtags further illustrate no shortage of public interest in ophthalmology, ocular surgery, and eye disease.

Given the significance of social media in the modern world, we recommend further studies to explore the potential of TikTok as a platform for ophthalmologists to promote the field and educate the general public on eye disorders and surgeries.

## Data Availability

All data produced in the present work are contained in the manuscript.

